# Advanced Multimodal AI for Predicting Long-Term Functional Outcomes After Ischemic Stroke Using Only Admission Data

**DOI:** 10.64898/2026.05.27.26354289

**Authors:** Fiona McBride, Haoxu Huang, Anjali Kiran Kapoor, Eric Karl Oermann, Jennifer Frontera, Narges Razavian

## Abstract

**Background and Purpose:** Prognostication after acute ischemic stroke often relies on limited variables and simple risk scores, despite richer information being available at admission. We developed a multimodal AI model using admission data to predict modified Rankin Scale (mRS) outcomes and compared it to established tools.

**Methods:** In a retrospective study of ischemic stroke/TIA patients, we trained three modality-specific models on admission non-contrast head CT, history and physical notes, and structured clinical variables, and combined them in a weighted-average ensemble. We predicted binary (mRS 0-2 versus 3-6) and ordinal mRS (0-6) outcomes at discharge and 90 days. Performance on an external test cohort was compared with THRIVE and SPAN-100 scores using AUROC, AUPRC, Brier score, mean absolute error (MAE), and quadratic weighted kappa (QWK).

**Results:** A total of 6,915 patients were split into training, validation and testing cohorts in a 3:1:1 ratio. For discharge binary mRS (n=1596), the multimodal ensemble achieved significantly better discrimination (AUROC 0.859, AUPRC 0.858) with 25-61% lower Brier scores than THRIVE or SPAN-100 (all p<0.001). For 90-day binary mRS (n=207), the model also outperformed both THRIVE and SPAN-100 (AUROC 0.838, AUPRC 0.805, with 3-38% lower Brier scores).

Ordinal mRS prediction showed similarly strong performance with significantly better QWK at discharge and numerically lower MAE. The multimodal ensemble model reassigned about one-third of patients to different risk categories versus THRIVE and was closer to the true discharge outcome in ∼74% of discordant cases.

**Conclusions:** We developed a well-calibrated multimodal AI model for prediction of discharge and 90-day post-stroke functional outcomes using only data present at the time of admission. This model outperforms existing prognostic tools and can support early clinical decision-making.

## Introduction

Ischemic stroke is a leading cause of long-term disability worldwide, and accurate early prediction of functional outcomes is critical for guiding treatment decisions and setting realistic recovery expectations for patients and families^1–4^. The first hour after emergency department presentation is often referred to as the “golden hour”. This is the timeframe when critical high impact decisions including thrombolysis, thrombectomy, surgical intervention, and triage are addressed. In severe circumstances, establishing goals of care may also be a priority early after presentation. Currently, prognostication is often based on expert clinician opinion or analog risk calculators. Existing functional outcome prediction scores such as THRIVE, and SPAN-100 rely on a limited set of variables and linear assumptions that underutilize the rich, multimodal information available at hospital admission ^2,3,5,6^.

In this project, we aimed to develop and externally validate a multimodal deep learning model built on foundational models that predicts both dichotomized and ordinal discharge and 90-day functional outcomes measured by the modified rankin scale (mRS)^3^ using data available at the time of admission. We integrate full volume 3D non-contrast admission head CT imaging, as well as structured and unstructured admission electronic health record (EHR) data and compare performance of our model to traditional predictive scores including THRIVE^5,7,8^ and SPAN-100^6^. Accurate and reliable outcome prediction utilizing admission imaging and clinical data could be used to inform early treatment decisions and shared decision making.

## Methods

### Study Design and Population

This is a retrospective cohort study utilizing adult ischemic stroke and transient ischemic stroke (TIA) patients enrolled in Get-With-The-Guidelines (GWTG) at a Joint Commission-certified Comprehensive Stroke Center in New York City (NYU Langone Hospital-Tisch and NYU Langone Hospital-Brooklyn) between January 2017 and February 2025. NYU Long Island operated as an independent Comprehensive Stroke Center until 2022, has clinicians and workflow protocols distinct from the other two campuses, and serves as an external test cohort for this study. Consecutive ischemic stroke patients were screened for inclusion in GWTG daily at all sites and all labelled GWTG data was manually collected or extracted from the EHR by trained stroke coordinators. Inclusion criteria were: age ≥18 years, acute ischemic stroke or TIA diagnosis, admission head CT DICOM available, admission history and physical (H&P) note available in the EHR (Epic), and discharge mRS and/or 90-day mRS documented **(Figure 1)**.

**Figure 1.**
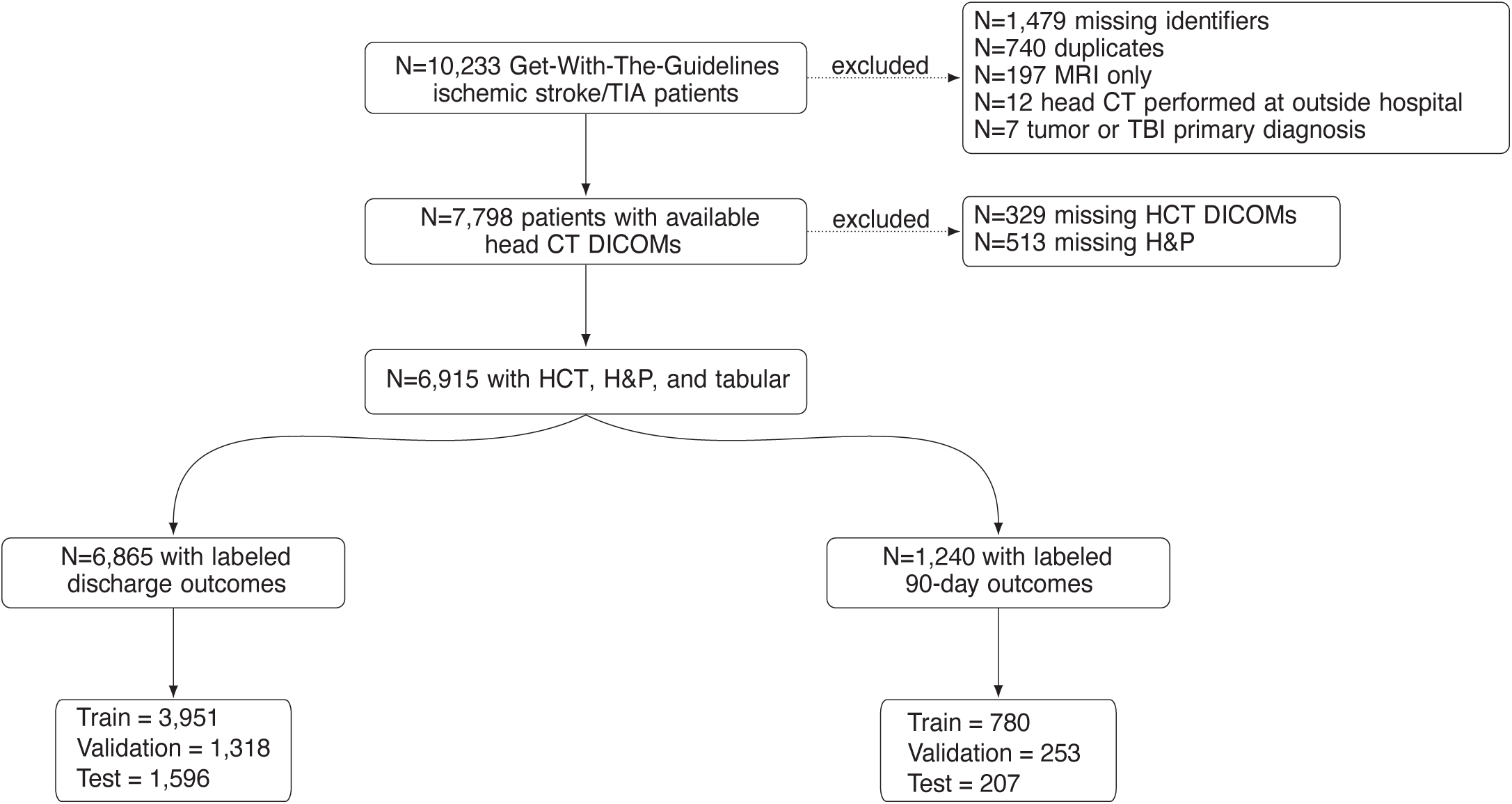
Inclusion and exclusion criteria.

Patients with hemorrhagic stroke (intracerebral hemorrhage or subarachnoid hemorrhage), stroke related to neurosurgical procedures, or primary diagnoses of intracranial tumor or traumatic brain injury were excluded **(Figure 1)**.

### Primary Outcomes

The co-primary outcomes of interest were discharge mRS and 90-day mRS. Per GWTG protocol, discharge mRS was collected in all patients by certified examiners and 90-day mRS was collected via phone interview with the patient or surrogate in those who received intravenous thrombolysis and/or endovascular treatment. No formal blinding of outcome assessment was performed or recorded. Accordingly, analyses of 90-day mRS are restricted to this treated subgroup and may not be generalizable to patients with stroke who did not receive these therapies. Both discharge and 90-day mRS were analyzed as: (1) an ordinal measure ranging from 0-6 and, (2) dichotomized as good (0-2) vs. poor (3-6) outcome.

### Cohort Partitioning

For each outcome of interest (discharge or 90-day mRS), the NYU Langone Hospitals (Tisch and Brooklyn) cohort was divided into training and internal validation sets. For discharge mRS, this yielded N=3,951 training and N=1,318 internal validation subjects. For 90-day mRS, this yielded N=780 training and N=253 internal validation subjects. Results are reported from the external test set from NYU Long Island, which included N=1,596 patients with discharge mRS and N=207 patients with 90-day mRS (**Figure 1**).

### Input Data and Preprocessing

Input data consisted of three modalities: 3D head CT DICOMs, unstructured admission H&P notes, and structured clinical variables. Non-contrast head CT scans were performed at admission on Siemens and Toshiba machines. We included 5 mm thickness scans with kVp values between 70-150 and convolution kernels Hr/Qr/J with sharpness levels of 35-45^9^. Non-contrast head CT scans obtained at admission were preprocessed by only preserving brain tissues (>-200 Hounsfield Units [HU]), removing background bias, and then setting all air regions to - 1024 HU. Then, three windowing levels (brain 40±40 HU, soft tissue 80±100 HU, bone 600±2800 HU) are applied on the head CT to construct the three channel scan. Admission H&P unstructured notes were extracted from the EHR via NYUMC Neuro Data Hub ^10^. H&P notes were preprocessed using MedGemma’s native tokenizer, which segments text into model-specific tokens (e.g., words or subwords) with a token limit of 8192. To mimic real-world deployment scenarios, no manual text selection or pre-processing was performed. The structured tabular clinical data consisted of admission variables including demographics (e.g., age, sex), baseline stroke severity measured by the NIH Stroke Scale (NIHSS)^11^, and past medical history (**Supplementary Table 1**).

### Model Architecture

We trained separate encoders for each data modality—non-contrast head CT, admission H&P notes, and structured clinical data—using pretrained foundation models^9,12,13^ as feature extractors (**Figure 2**).

**Figure 2.**
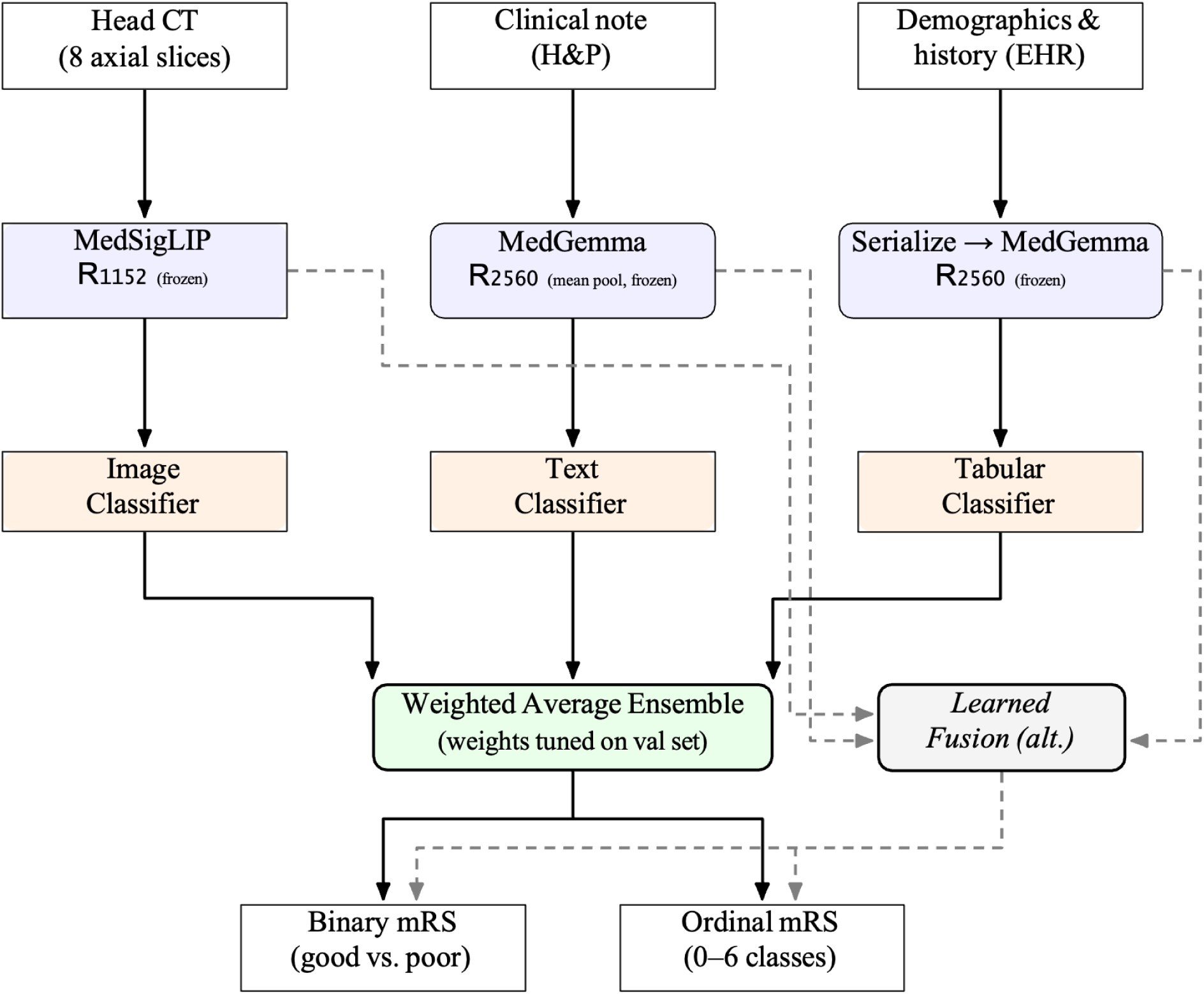
Model architecture. Head CT images, clinical notes (H&P), and demographics/history from the EHR are processed through modality-specific foundation models—FM-HCT for imaging, MedGemma for text, and TabPFN for tabular data—followed by separate image, text, and tabular classifiers. The resulting modality-specific predictions are combined using a weighted average ensemble, with ensemble weights tuned on the validation set, to generate both binary mRS (good vs. poor) and ordinal mRS (0–6 classes) outputs. Dashed arrows indicate an alternative learned-fusion strategy and optional cross-modal feature integration explored in addition to the primary ensemble pipeline. FM=foundation model; EHR=electronic health record; H&P=history and physical; alt.=alternate; val=validation; mRS=modified Rankin Score

**CT encoder.** Head CT scans were processed with the NYU Head CT Foundation Model, a 3D Vision Transformer (ViT-Base) pretrained on 361,663 head CT scans^9^. Input volumes were downsampled to 96×96×96 voxels and partitioned into 12×12×12 patches, yielding 512 tokens per scan. The CT encoder was initialized from pretrained weights and fine-tuned on the study cohort.

**Text encoder.** Unstructured admission H&P notes were encoded using MedGemma^12^, a 4-billion-parameter multimodal vision–language model trained on medical data. While MedGemma has both text and imaging capabilities, we used only the text encoder because the vision encoder does not support 3D imaging. Notes were tokenized with MedGemma’s native tokenizer (context window 8,192 tokens). The median token length for H&P notes was 2,414 (IQR 1,805-3,266). Text embeddings were precomputed and kept frozen during multimodal training.

**Tabular encoder.** Structured clinical features were processed with TabPFN^13^, a foundation model for tabular data. Rather than using TabPFN’s classification outputs, we extracted internal representations via the TabPFNEmbedding interface, using 5-fold cross-fitting on the training set to avoid leakage. This produced a 192-dimensional embedding per patient, which was then held fixed and used as the tabular input to downstream fusion models.

The head CT encoder was fine-tuned end-to-end using a reduced learning rate (1×10^-5^) to adapt pretrained CT representations to stroke outcome prediction, while text and tabular embeddings were precomputed from frozen encoders prior to training to reduce GPU memory requirements during fine-tuning.

### Model Training, Validation and Testing

All models were trained separately for discharge and 90-day mRS prediction. For each outcome, we trained a CT-only model, a text-only model, a tabular-only model, and multimodal fusion models that combined the three encoders.

For ordinal mRS (0–6), models output a 7-dimensional probability distribution over mRS categories and were trained with a soft ordinal cross-entropy loss that penalizes larger deviations more heavily. The predicted mRS used for MAE and correlation metrics was the expected value of this distribution. For the binary outcome (good [0–2] vs poor [3–6]), models output a scalar probability and were trained with binary cross-entropy loss.

Optimization used AdamW^14^ with cosine learning rate decay and mixed-precision training. We used a batch size of 64 and trained for 5 epochs per configuration. Hyperparameters were selected based on performance on the internal validation set. All models used the fixed training/validation/test splits described above; performance is reported on the external test cohort.

### Model Fusion

The primary multimodal model was a weighted-average ensemble. For each patient and outcome, the CT, text, and tabular encoders each produced a probability distribution over mRS categories (ordinal 0–6 or binary 0–2 vs 3–6). These modality-specific predictions were combined as a convex combination, with modality weights optimized on the internal validation set. This scheme introduces no additional trainable parameters and remains operational when one or more modalities are missing.

To contextualize the ensemble, we also evaluated alternative fusion strategies. We tested late fusion, where the final embedding from each individual encoder is projected to a common 512-dimensional representation, and the resulting vectors are concatenated and passed through a two-layer multilayer perceptron which outputs a single prediction. We also tested intermediate fusion, in which tabular and text embeddings are projected into the vision transformer’s token space and injected into the image encoder at an intermediate layer (layer 6 of 12), allowing subsequent self-attention layers to process imaging and clinical features jointly before producing a final prediction. The weighted-average ensemble was selected as the primary model based on its external test performance, simplicity, and robustness to missing modalities.

To quantify modality contributions, we report performance of the unimodal encoders (CT-only, text-only, tabular-only) trained with the same splits, loss functions, and optimization scheme as the ensemble, and compare them directly to the multimodal ensemble on the external test set.

### Evaluation Metrics

For the binary mRS outcome (good [0–2] vs poor [3–6]), model performance was evaluated in terms of both discrimination and calibration. Discrimination was assessed using the area under the receiver operating characteristic curve (AUROC) and the area under the precision–recall curve (AUPRC)^15,16^.

Calibration of the binary predictions was quantified using the Brier score. The Brier score is the mean squared error between the predicted probability and the observed binary outcome (0 for good outcome, 1 for poor outcome), with lower values indicating better overall calibration and accuracy of the probabilistic predictions. Calibration curves were generated using reliability diagrams with quantile binning with 10 bins of equal sample size, plotting mean predicted probability against observed outcome frequency for each model. THRIVE and SPAN-100 scores were linearly rescaled to [0,1] prior to plotting, as these are ordinal risk scores rather than calibrated probabilities.

For the ordinal mRS outcome (0–6 scale), we evaluated model performance using metrics that capture both categorical agreement and rank-based concordance. Agreement between predicted and observed mRS categories was measured with the quadratic weighted kappa (QWK), which weights disagreements more heavily when the predicted and true categories are farther apart^17^. QWK ranges from 0 to 1, with values closer to 1 indicating higher agreement.

QWK values of 0.60-0.80 generally indicate good agreement, while values >0.80 indicate excellent agreement^18^. We also report the mean absolute error (MAE) between the predicted and observed mRS, computed as the average absolute difference between the predicted mRS (taken as the expectation over the predicted probability distribution) and the true mRS value.

Spearman’s rank correlation coefficient was used to measure the correlation between the model’s predicted mRS and the true observed mRS. We calculated Earth Mover’s Distance (EMD) to measure the similarity between our predicted mRS distribution and the true mRS distribution, with a lower EMD representing greater similarity.

For each outcome formulation, AUROC and AUPRC (binary mRS) and QWK and MAE (ordinal mRS) were prespecified as primary performance metrics; Spearman correlation and EMD were considered supportive.

### Comparator Metrics

As clinical comparators, we evaluated the THRIVE^5^ and SPAN-100^6^ scores. The THRIVE score is a validated ischemic stroke risk score based on age, NIH Stroke Scale (NIHSS), and comorbidities (hypertension, diabetes mellitus, atrial fibrillation), with total scores ranging from 0 to 9. Higher scores indicate a higher likelihood of poor functional outcome (mRS 3-6) at 90-days. The SPAN-100 index is defined as the sum of age and NIHSS; patients with SPAN-100 ≥100 are classified as high risk for poor outcome at 90-days (mRS 3-6). For both scores, we computed predicted probabilities of poor outcome using their published categorizations and used these as reference models for comparison.

### Statistical Comparison

For all metrics, we compared the multimodal ensemble to established clinical outcome scores (THRIVE and SPAN-100), as well as to single-modality models. AUROCs for paired models were compared using the DeLong test^19^. Differences in AUPRC, Brier score, QWK, MAE, and Spearman correlation were estimated along with 95% confidence intervals using bootstrap resampling-based procedures. P-values were adjusted for multiple comparisons (across metrics and comparator models) using the Bonferroni correction, and corrected P-values are reported. Recalibration specificity and sensitivity were compared using McNemar’s test because we were comparing paired data in a 2×2 contingency table^20^. Unless otherwise specified, all reported metrics are computed on the external test set. Statistical tests were run in Python (3.6.25).

### Code and Reproducibility

This study was approved by the NYU Institutional Review Board with waiver of informed consent. The study was reported in accordance with the Transparent Reporting of Multivariable Prediction Model for Individual Prognosis or Diagnosis using Artificial Intelligence (TRIPOD+AI) guidelines. Code is available at github.com/mcbride-fiona/Multimodal_AI_for_Stroke_Outcomes. The corresponding authors had full access to all study data and take responsibility for the integrity of the data and the accuracy of the data analysis. No formal study protocol was prepared, and this study was not registered. The study data are not publicly available because they contain protected health information. Patients and the public were not involved in the design, conduct, reporting, interpretation, or dissemination of this study.

## Results

### Cohort Characteristics

The initial cohort included 10,233 Get With The Guidelines (GWTG) ischemic stroke or transient ischemic attack (TIA) encounters^21^. After applying exclusion criteria, 6,915 ischemic stroke/TIA encounters with complete admission-level data across all three modalities remained (**Figure 1**). For discharge mRS, encounters were assigned in a 3:1:1 ratio to fixed training (n=3,951), internal validation (n=1,318), and held-out external test (n=1,596) sets. For 90-day mRS, encounters were also assigned in a 3:1:1 ratio to fixed training (n=780), internal validation (n=253), and held-out external test (n=207) sets. The external test cohort was significantly older, had lower admission NIHSS, higher prevalence of coronary artery disease, carotid stenosis, diabetes, obesity, hyperlipidemia, renal insufficiency, and dementia, and lower prevalence of drug/alcohol use, and tobacco use (**Table 1**). Notably, baseline mRS did not differ across groups.

**Table 1:** Characteristics of patients in training, testing and validation cohorts. Statistically significant differences between sites were observed for several variables, reflecting the external site’s distinct case mix; however, most absolute differences were small and consistent with expected multi-site variation.

| Variable | Discharge<br>mRS<br>Training | Discharge<br>mRS<br>Internal<br>Validation | Discharge<br>mRS<br>External<br>Testing | 90d mRS<br>Training | 90d mRS<br>Internal<br>Validation | 90d mRS<br>External<br>Testing |
| --- | --- | --- | --- | --- | --- | --- |
| N | 3951 | 1318 | 1596 | 780 | 253 | 207 |
| **Demographics** |  |  |  |  |  |  |
| Age (years),<br>median (IQR) | 71 (60–81) | 71 (60–82) | 74 (62–83)* | 73 (62–82) | 71 (57–82) | 77 (64–85)* |
| Sex (female),<br>N (%) | 1855 (47.0%) | 628 (47.6%) | 788 (49.4%) | 391 (50.1%) | 119 (47.0%) | 97 (46.9%) |
| Baseline<br>mRS, median<br>(IQR) | 0 (0–2) | 0 (0–2) | 0 (0–2) | 0 (0–2) | 0 (0–1) | 0 (0–1) |
| Admission<br>NIHSS,<br>median (IQR) | 3 (1–8) | 3 (1–8) | 2 (0–6)* | 8 (4–20) | 11 (3–21) | 10 (4–16) |
| **Past Medical History, N (%)** |  |  |  |  |  |  |
| Atrial<br>fibrillation | 681 (17.2%) | 210 (15.9%) | 272 (17.0%) | 163 (20.9%) | 54 (21.3%) | 39 (18.8%) |
| Prosthetic<br>heart valve | 31 (0.8%) | 2 (0.2%) | 4 (0.3%) | 5 (0.6%) | 0 (0.0%) | 0 (0.0%) |
| Coronary<br>artery disease | 734 (18.6%) | 268 (20.3%) | 386 (24.2%)* | 154 (19.7%) | 51 (20.2%) | 46 (22.2%) |
| Carotid<br>stenosis | 80 (2.0%) | 32 (2.4%) | 53 (3.3%)* | 17 (2.2%) | 4 (1.6%) | 4 (1.9%) |
| Diabetes | 1262 (31.9%) | 424 (32.2%) | 556 (34.8%)* | 207 (26.5%) | 63 (24.9%) | 59 (28.5%) |
| Hypertension | 2816 (71.3%) | 953 (72.3%) | 1140 (71.4%) | 540 (69.2%) | 177 (70.0%) | 130 (62.8%) |
| Peripheral<br>vascular<br>disease | 96 (2.4%) | 31 (2.4%) | 33 (2.1%) | 20 (2.6%) | 3 (1.2%) | 1 (0.5%) |
| Tobacco use | 485 (12.3%) | 157 (11.9%) | 65 (4.1%)* | 78 (10.0%) | 29 (11.5%) | 8 (3.9%)* |
| Hyperlipidem<br>ia | 1873 (47.4%) | 634 (48.1%) | 807 (50.6%)* | 346 (44.4%) | 104 (41.1%) | 89 (43.0%) |
| **Heart Failure** | 253 (6.4%) | 108 (8.2%) | 132 (8.3%) | 50 (6.4%) | 23 (9.1%) | 16 (7.7%) |
| **Prior stroke/TIA** | 975 (24.7%) | 324 (24.6%) | 403 (25.3%) | 158 (20.3%) | 51 (20.2%) | 42 (20.3%) |
| **Drug/alcohol use** | 131 (3.3%) | 52 (3.9%) | 26 (1.6%)* | 26 (3.3%) | 11 (4.3%) | 4 (1.9%) |
| **Obesity** | 163 (4.1%) | 52 (3.9%) | 101 (6.3%)* | 26 (3.3%) | 12 (4.7%) | 11 (5.3%) |
| **Renal insufficiency** | 244 (6.2%) | 101 (7.7%) | 144 (9.0%)* | 36 (4.6%) | 16 (6.3%) | 14 (6.8%) |
| **Sleep apnea** | 100 (2.5%) | 34 (2.6%) | 49 (3.1%) | 16 (2.1%) | 4 (1.6%) | 1 (0.5%) |
| **Depression** | 186 (4.7%) | 57 (4.3%) | 74 (4.6%) | 37 (4.7%) | 9 (3.6%) | 8 (3.9%) |
| **Venous thromboembolism** | 126 (3.2%) | 35 (2.7%) | 63 (3.9%) | 23 (2.9%) | 5 (2.0%) | 6 (2.9%) |
| **Dementia** | 102 (2.6%) | 31 (2.4%) | 73 (4.6%)* | 21 (2.7%) | 5 (2.0%) | 9 (4.3%) |
| **Cancer** | 24 (0.6%) | 10 (0.8%) | 12 (0.8%) | 5 (0.6%) | 2 (0.8%) | 0 (0.0%) |
| **Acute Stroke Treatment, N (%)** |  |  |  |  |  |  |
| **IV thrombolysis** | 527 (13.3%) | 159 (12.1%) | 161 (10.1%)* | 404 (51.8%) | 123 (48.6%) | 135 (65.2%)* |
| **Mechanical thrombectomy** | 602 (15.2%) | 215 (16.3%) | 109 (6.8%)* | 439 (56.3%) | 155 (61.3%) | 84 (40.6%)* |
| **Outcomes, N (%)** |  |  |  |  |  |  |
| **mRS 0** | 681 (17.2%) | 233 (17.7%) | 429 (26.9%)* | 179 (22.9%) | 62 (24.5%) | 67 (32.4%)* |
| **mRS 1** | 900 (22.8%) | 295 (22.4%) | 289 (18.1%)* | 137 (17.6%) | 36 (14.2%) | 31 (15.0%) |
| **mRS 2** | 489 (12.4%) | 164 (12.4%) | 112 (7.0%)* | 89 (11.4%) | 34 (13.4%) | 21 (10.1%) |
| **mRS 3** | 486 (12.3%) | 180 (13.7%) | 166 (10.4%)* | 66 (8.5%) | 16 (6.3%) | 22 (10.6%) |
| **mRS 4** | 1014 (25.7%) | 305 (23.1%) | 459 (28.8%)* | 107 (13.7%) | 24 (9.5%) | 21 (10.1%) |
| **mRS 5** | 308 (7.8%) | 110 (8.3%) | 122 (7.6%) | 33 (4.2%) | 14 (5.5%) | 8 (3.9%) |
| **mRS 6** | 73 (1.8%) | 31 (2.4%) | 19 (1.2%) | 169 (21.7%) | 67 (26.5%) | 37 (17.9%) |
| **mRS 0–2** | 2070 (52.4%) | 692 (52.5%) | 830 (52.0%) | 405 (51.9%) | 132 (52.2%) | 119 (57.5%) |
| **mRS 3–6** | 1881 (47.6%) | 626 (47.5%) | 766 (48.0%) | 375 (48.1%) | 121 (47.8%) | 88 (42.5%)* |

\*p<0.05 training and validation vs. external testing for that outcome cohort

### Selection of Model Fusion Method

We tested alternate methods of combining the signals from the individual data encoders. Late fusion outperformed intermediate fusion on binary discharge mRS (AUROC 0.848 vs. 0.830), binary 90-day mRS (AUROC 0.847 vs. 0.793), ordinal discharge mRS (AUROC 0.757 vs.

0.744), and ordinal 90-day mRS (AUROC 0.671 vs. 0.611). However, the weighted average ensemble of all modality combinations achieved the highest performance for discharge mRS (binary AUROC 0.859; ordinal AUROC 0.761) and comparable performance for 90-day mRS (binary AUROC 0.838; ordinal AUROC 0.667). Since ensemble fusion requires less peak GPU memory during training it was used as the primary model for all analyses **(Supplementary Figure 1)**.

### Binary mRS Outcomes Compared to Traditional Models

**Discharge mRS.** On the external test set (n=1,596; 766 poor outcomes, 830 good outcomes), the multimodal weighted-average ensemble model achieved robust discrimination and calibration with AUROC 0.859 (95% CI 0.840–0.877), AUPRC 0.858 (95% CI 0.835–0.879), and a Brier score of 0.156 **(Figure 3**). Compared with THRIVE, the ensemble showed absolute improvements of 9.3% in AUROC (vs. 0.766, p<0.001) and 11.5% in AUPRC (vs. 0.743, p<0.001), with a 25% lower Brier score (vs. 0.207, p<0.001). Compared with SPAN-100, improvements were 27.4% in AUROC (vs. 0.585, p<0.001), 29.4% in AUPRC (vs. 0.565, p<0.001), and 61% in Brier score (vs. 0.399, p<0.001) (**Table 2**).

**Figure 3:**
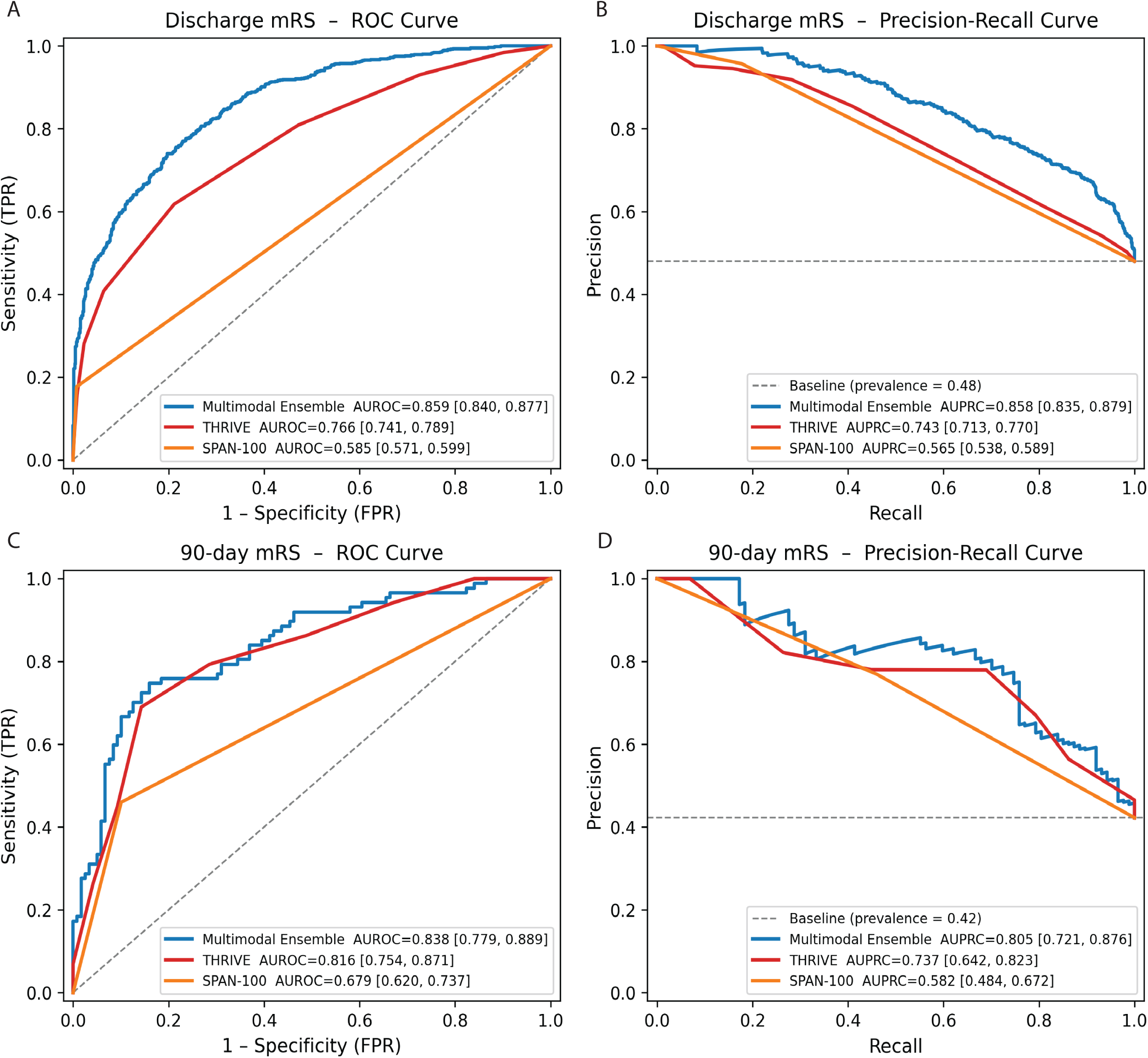
AUROC curves for discharge (A) and 90-day (C) binary mRS (0-2 versus 3-6) outcomes and AUPRC curves for discharge (B) and 90-days (D) binary mRS (0-2 versus 3-6) outcomes for the multimodal weighted average ensemble compared to THRIVE and SPAN-100. The multimodal ensemble model had significantly better AUROC and AUPRC for discharge mRS, and AUPRC for 90-day mRS compared to both THRIVE and SPAN-100 (all P<0.001). The multimodal ensemble also performed significantly better than SPAN-100 for 90-day AUROC, and demonstrated a non-significant improvement in 90-day AUROC compared to THRIVE.

**Table 2:** Binary mRS Prediction Performance on the External Test Set. Discrimination (AUROC, AUPRC) and calibration (Brier score) for the multimodal weighted-average ensemble versus THRIVE and SPAN-100 at discharge (n=1,596) and 90 days (n=207). Metrics are reported with 95% confidence intervals.

| Outcome | Model | N | AUROC (95% CI) | AUPRC (95% CI) | Brier score (95% CI) |
| --- | --- | --- | --- | --- | --- |
| Discharge mRS | Ensemble | 1596 | 0.859 (0.840–0.877)*<sup>+</sup> | 0.858 (0.835–0.879)*<sup>+</sup> | 0.156 (0.148–0.165)*<sup>+</sup> |
| Discharge mRS | THRIVE | 1596 | 0.766 (0.742–0.789) | 0.743 (0.713–0.770) | 0.207 (0.197–0.217) |
| Discharge mRS | SPAN-100 | 1596 | 0.585 (0.571–0.599) | 0.565 (0.538–0.589) | 0.399 (0.375–0.424) |
| 90-day mRS | Ensemble | 207 | 0.838 (0.779–0.889)<sup>+</sup> | 0.805 (0.721–0.876)*<sup>+</sup> | 0.176 (0.158–0.195)<sup>+</sup> |
| 90-day mRS | THRIVE | 207 | 0.816 (0.754–0.871) | 0.737 (0.642–0.823) | 0.179 (0.155–0.207) |
| 90-day mRS | SPAN-100 | 207 | 0.679 (0.620–0.737) | 0.582 (0.484–0.672) | 0.286 (0.223–0.350) |

\* Ensemble model significantly better ( $p<0.05$ ) compared to THRIVE, <sup>+</sup> Ensemble model significantly better ( $p<0.05$ ) compared to SPAN-100

**90-Day mRS.** For 90-day prediction (n=207; 88 poor outcomes, 119 good outcomes), the ensemble model achieved AUROC 0.838 (95% CI 0.779–0.889) and AUPRC 0.805 (95% CI 0.721–0.876), with a Brier score of 0.176 **(Figure 3)**. The ensemble significantly outperformed SPAN-100 (AUROC improvement 15.8%, p<0.001; AUPRC improvement 22.4%, p<0.001; Brier score 0.176 vs. 0.286, p<0.001, **(Table 2**), and demonstrated significant improvement in AUPRC (9.23%, P=0.015) compared to THRIVE, with non-significant improvement in AUROC (2.67%, P=0.302), and Brier score (1.68%, P=0.736) compared to THRIVE.

Calibration plots (**Figure 4**) demonstrate that the ensemble model generated predicted probabilities of poor outcome at discharge and 90-days that more closely approximated the actual observed outcomes than either THRIVE or SPAN-100, indicating more reliable absolute probability estimates.

**Figure 4.**
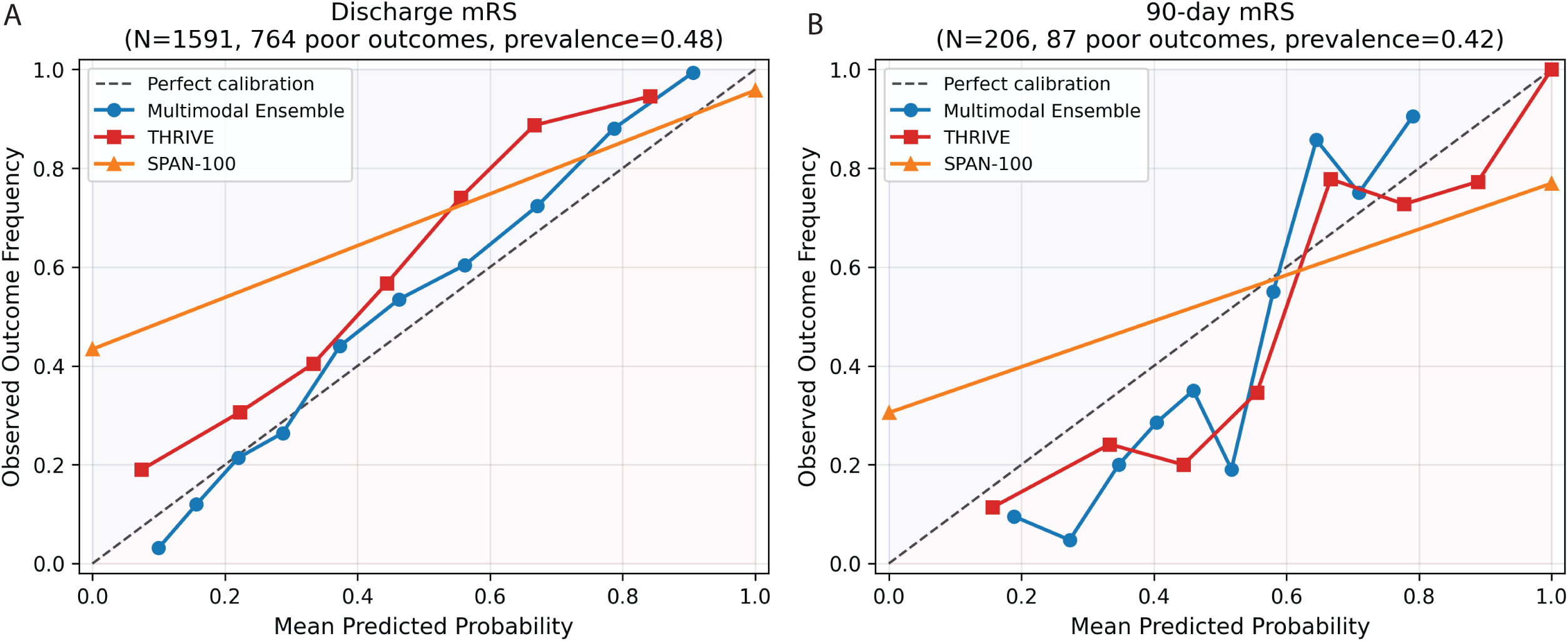
Calibration diagrams compare observed outcome frequency versus mean predicted probability for the multimodal ensemble, THRIVE, and SPAN-100 models for predicting poor functional outcome. (A) shows calibration for discharge mRS in the full cohort, and (B) shows calibration for 90-day mRS. The dashed diagonal indicates perfect calibration. Across both endpoints, the multimodal ensemble generally tracks the ideal calibration line more closely than the comparator clinical scores, suggesting improved probability calibration for predicting poor outcomes.

### Ordinal mRS Outcomes (0–6) Compared to Traditional Models

**Discharge mRS.** The ensemble model achieved an average AUROC of 0.761 (95% CI 0.742–0.776) across all 7 levels of the mRS, as well as QWK 0.613 (95% CI 0.577–0.647), MAE 1.061 (95% CI 1.003–1.119), EMD 1.520 (95% CI 1.494-1.546), and Spearman ρ between the predicted and true mRS labels of 0.684 (95% CI 0.652–0.712), significantly outperforming THRIVE across all ordinal metrics (all p≤0.001, **Table 3**). The ensemble model was 73.6% accurate (95% CI 71.5–75.9%) at predicting discharge mRS within 1 level of the actual outcome, significantly better than THRIVE (68.6%, p=0.001). The ensemble model performed better at predicting extreme mRS levels (mRS 0 AUROC: 0.85; mRS 5–6: 0.89 and 0.85) than intermediate scores (mRS 1–4 AUROC: 0.65–0.75) (**Figure 5**). Conversely, AUPRC measures were generally poor for both the ensemble model and THRIVE across many levels of the mRS, reflecting the low prevalence of certain outcomes in the test data. Ordinal metrics were not compared against SPAN-100 due to the binary nature of the SPAN-100 prediction scale.

**Figure 5:**
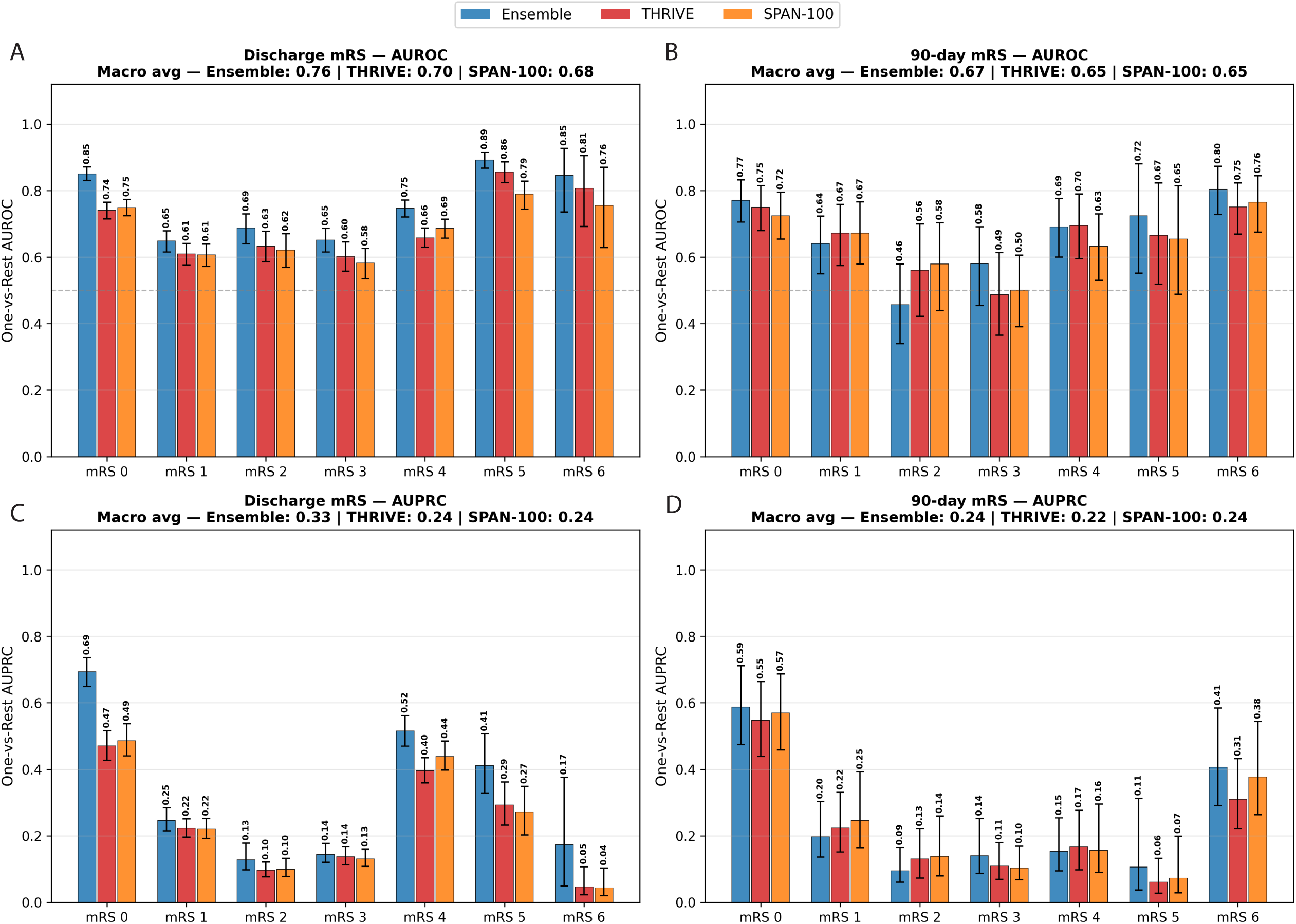
Per-Class AUROC and AUPRC for Ordinal mRS Prediction at Discharge and 90 Days. One-vs-rest AUROC for each mRS level (0–6) on the external test set (NYU Long Island) at discharge (A; n=1,596) and 90 days (B; n=207). One-vs-rest AUPRC for each mRS level (0–6) on the external test set (NYU Long Island) at discharge (C; n=1,596) and 90 days (D; n=207). Error bars represent 95% bootstrap confidence intervals. Both timepoints show strongest discrimination at the outcome extremes (mRS 0, 5–6) and weakest performance at intermediate disability levels (mRS 1–4). At discharge, the ensemble model had significantly higher macro AUROC and AUPRC compared to both THRIVE and SPAN-100 (all p<0.001). At 90 days, the ensemble had significantly higher AUPRC compared to THRIVE (p=0.033) but not SPAN-100. The macro AUROC at 90 days was not significantly different between any of the models.

**Table 3:** Ordinal mRS Prediction Performance on the External Test Set. Agreement (QWK, Within±1 accuracy), rank correlation (Spearman ρ), error (MAE), and distributional similarity (EMD) for the multimodal weighted-average ensemble versus THRIVE across all 7 mRS levels (0–6) at discharge (n=1,596) and 90 days (n=207). Metrics are reported with 95% confidence intervals estimated by bootstrap resampling. SPAN-100 was excluded from ordinal comparisons due to its binary prediction scale.

| Outcome | Model | N | QWK (95% CI) | MAE (95% CI) | Spearman (95% CI) | EMD (95% CI) | Within$\pm 1$ Accuracy (95% CI) |
| --- | --- | --- | --- | --- | --- | --- | --- |
| Discharge mRS | Ensemble | 1596 | 0.613 (0.577–0.647)* | 1.061 (1.003–1.119)* | 0.684 (0.652–0.712)* | 1.520 (1.494–1.546)* | 0.736 (0.715–0.759)* |
| Discharge mRS | THRIVE | 1596 | 0.461 (0.422–0.501) | 1.263 (1.201–1.327) | 0.519 (0.479–0.557) | 1.624 (1.597–1.651) | 0.686 (0.663–0.708) |
| 90-day mRS | Ensemble | 207 | 0.554 (0.443–0.649) | 1.607 (1.379–1.850) | 0.608 (0.514–0.683)* | 2.169 (2.101–2.240) | 0.592 (0.524–0.665) |
| 90-day mRS | THRIVE | 207 | 0.476 (0.371–0.575) | 1.811 (1.551–2.102) | 0.548 (0.444–0.638) | 2.006 (1.903–2.115) | 0.558 (0.490–0.626) |

\*Ensemble model significantly better ( $p<0.05$ ) compared to THRIVE

**90-Day mRS.** The ensemble achieved an average AUROC of 0.667 (95% CI 0.624–0.707) across the 7 levels of the mRS, with a significantly better Spearman ρ 0.612 (95% CI 0.520–0.691) compared to THRIVE (all p<0.05). The ensemble model MAE and QWK were numerically better than THRIVE but did not reach statistical significance. The EMD of the ensemble was slightly worse than that of THRIVE. The ensemble model was 59.2% accurate (95% CI 52.4–66.5%) at predicting 90-day mRS within 1 level of the actual outcome, while THRIVE was 55.8% accurate (**Table 3**). As with the discharge ordinal mRS prediction, the ensemble model performed better at predicting mRS 0, 5, and 6, with AUROCs of 0.773, 0.722, and 0.808 respectively, while AUROCs for intermediate grades (mRS 1–4) were mediocre (0.46–0.69, **Figure 5**). AUPRC values were poor for both the ensemble model and THRIVE across most levels of the mRS.

### Reclassification Analysis Against THRIVE

**Discharge mRS.** The weighted-average ensemble and THRIVE assigned patients to different risk categories in 35% of encounters (κ=0.55). True crossover reclassifications where THRIVE predicted a very high mRS and our model predicted a very low mRS, or vice versa, were rare (≤1.3%); the dominant disagreements were adjacent-level shifts **(Supplementary Figure 2)**. In discordant encounters, the ensemble was closer to the true outcome in 73.5% of cases compared to THRIVE. At the individual mRS level, the ensemble model showed an advantage at the outcome extremes: for patients with observed mRS 0 (no disability), the ensemble predicted the correct category in 64.8% of cases versus 36.8% for THRIVE, and for observed mRS 5 (severe disability), the ensemble predicted the correct category in 45.9% versus 0.8% with THRIVE. THRIVE was modestly more accurate for intermediate categories mRS 1 (57.1% vs. 45.3%) and mRS 4 (56.0% vs. 38.3%), reflecting the alignment of its binary comorbidity inputs with mid-range disability **(Supplementary Figure 3)**. Both approaches showed low exact-match accuracy for mRS 2–3, though within-1 accuracy remained at 48–62%.

The ensemble model significantly improved sensitivity relative to THRIVE (48.4% vs. 28.1%, p<0.001) at the cost of a modest reduction in specificity (94.9% vs. 97.7%, p<0.001), reflecting a recalibration toward detection of true poor outcomes rather than conservative under-prediction. Notably, THRIVE classified only 14.7% of patients into the high-risk category (n=234) compared to 25.9% for the ensemble model (n=413), despite an observed poor outcome rate of 48.0%—suggesting that THRIVE systematically under-flags patients at discharge who go on to experience significant disability **(Supplementary Table 2)**.

**90-Day mRS.** Discordance between ensemble model and THRIVE predictions occurred in 32% of the 207 test encounters (κ=0.59). While the ensemble model did not demonstrate significantly greater accuracy in discordant 90-day encounters (45.5%, p=0.805), it showed improved specificity compared to THRIVE (93.3% vs. 85.7%, p=0.023), driven primarily by a reduction in over-classification of patients into the high-risk category (22.7% vs. 37.2%).

THRIVE classified 37.2% of patients as high risk despite an observed poor outcome rate of 42.5% and a PPV of only 77.9%, meaning approximately 1 in 5 patients flagged as high risk by THRIVE did not experience a poor outcome. The ensemble model’s higher PPV (83.0%) and lower high-risk classification rate (22.7%) reflect a recalibration away from systematic prognostic pessimism **(Supplementary Table 2)**. The lack of a significant discordant-case accuracy advantage likely reflects the bimodal distribution of 90-day outcomes in this treated cohort and the small number of patients at intermediate mRS grades in the external test set (n=8–22 per class), which limits class-level resolution **(Supplementary Figures 2 and 3)**.

At the class level, both the ensemble model and THRIVE identified mRS 0 with reasonable accuracy (ensemble: 64.2%, THRIVE: 59.7%) and mRS 6 with good accuracy (ensemble: 75.7%, THRIVE: 86.5%; **Supplementary Figure 3**). Neither approach correctly classified any patient with intermediate 90-day outcomes (mRS 2–4: 0% exact accuracy for both), consistent with the bimodal distribution of 90-day outcomes in this treated cohort and the small number of patients at intermediate mRS grades in the external test set (n=8–22 per class) **(Supplementary Figure 3)**.

## Discussion

In this study, we developed a multimodal AI ensemble model with strong discrimination metrics that produced robust discharge and 90-day functional outcome estimates using only data present at admission. Developing robust predictive models that can be deployed at the time of admission is important for many disease models where treatment decisions are time sensitive, acute deterioration is possible, and new therapeutic interventions are emerging. Our model outperformed traditional predictive scoring systems (THRIVE, SPAN-100) on most metrics for both dichotomized and ordinal mRS prediction at discharge and 90-days. Importantly, our ensemble model produced well-calibrated risk estimates while preserving strong discrimination and accurate rank ordering. This means a clinician using this tool can communicate to a patient’s family not only that the patient is at “high risk” but can provide a specific, trustworthy probability of achieving a given functional state—a capability that existing scoring tools do not reliably support. We found disagreement between our model and THRIVE outcome prediction in ∼30% of cases and our model was closer to the true discharge outcome in nearly three-quarters of discordant encounters, reclassifying the intermediate-risk cases where THRIVE provides the least actionable guidance. For 90-day outcomes, the THRIVE score tended to over-predict poor outcomes—reflected in its lower specificity. Over-classification of patients as high risk for poor outcome can lead to premature limitation of care and inappropriate prognostic pessimism. Our model improved specificity, even at the cost of modestly lower sensitivity, representing a meaningful recalibration that helps guide decision making.

Major strengths of our model are: 1) its use of admission only data, allowing for prognostic information early in the hospital course when high-stakes decision making occurs, 2) its generation of both dichotomized and ordinal mRS outputs including granular details from each level of the mRS that can be useful for shared decision making, 3) its generalizability since the data inputs (head CT and admission note) are standardly collected at stroke presentation, 4) the model is low touch and pragmatic; it does not require manual input of structured data and can be fully automated to rapidly produce outcome probability predictions, 5) its use of whole brain 3D head CT data, which does not require derived CT metrics or segmentation, 6) its use of foundation models, including open-source MedGemma, allowing for generation of fine-tuned models with less labelled data, less compute, and ultimately lower cost, and 7) the ensemble fusion design, which remains robust to missing data, and is more efficient than earlier fusion techniques that require more compute. Since each modality model is trained independently, ensemble fusion allows individual modality models to be updated or replaced without retraining the full system.

By contrast, prior studies that utilized AI-modeling to predict mRS post-stroke outcomes required up to 3-7 days of data to generate predictions^22,23^ utilized manually collected structured data, which is both time consuming and impractical for real-world implementation^5,22,24–26^, required imaging segmentation or extracted features, which add expense and additional software^24,25^, and/or required MRI, which is not typically available at the time of admission^22,24^. Furthermore, most studies focused on predicting dichotomized mRS outcomes^23–26^ without the capability of predicting meaningful differences captured by intermediate mRS scores.

Several limitations must be acknowledged. First, though our model performed well on discharge and 90-day dichotomized outcomes compared to traditional models, the performance for ordinal mRS prediction was less robust, particularly for intermediate levels of the mRS. This is, in part, related to the low prevalence of certain mRS outcomes and the fact that the dataset is modest in size relative to many machine learning benchmarks, particularly for labeled 90-day outcomes, limiting precision and statistical power. Nonetheless, there are a paucity of studies demonstrating reliable prediction of ordinal mRS outcomes at each level of the scale at 90-days, thus our study represents a meaningful contribution to the literature. The limitations in predicting individual mRS outcomes suggests that larger heterogeneous cohorts are needed for model training. Another limitation is that external validation was performed within the same health system, and generalizability to other systems, regions, and populations remains to be established. We did not formally assess model fairness or subgroup performance across sociodemographic groups, and therefore cannot exclude the possibility of differential performance across patient populations. Last, the ensemble model’s reliance on LLM-extracted text features introduces dependence on the specific language model and documentation style.

Further testing and external validation is warranted.

## Future Work

Causal inference and counterfactual prediction analyses can be constructed using our model to identify modifiable risk factors or potential interventions that could modulate outcome probabilities. Clinician-facing model-generated prognostic estimates can serve to democratize the delivery of stroke care, since current prognostication strategies rely heavily on clinician expertise and experience. Last, our prognostic model could improve hospital efficiency, triage, length of stay, observed to expected mortality, and 30-day readmission rates, all of which impact hospital reimbursement.

## Conclusions

In this retrospective cohort of ischemic stroke and TIA patients at a large academic health system, a multimodal ensemble model that integrates routinely collected admission CT, H&P notes, and structured clinical data generated more accurate and better-calibrated discharge and 90-day mRS predictions than established scores such as THRIVE and SPAN-100.

## Data Availability

The study data are not publicly available because they contain protected health information.

## Acknowledgments

FM, JAF, and NR are supported by NYU Langone Institute for Translational Neuroscience. NR and HH are also supported by NIH 5R01AG085617.

## Conflicts of Interest

FM, HH, AKK, JAF, and NR declare no conflicts of interest relevant to this article. EKO reports equity holdings in Artisight, MarchAI, and Eikon Therapeutics.

## Notes

### Author Declarations

This study was approved by the NYU Institutional Review Board with waiver of informed consent.

